# A Randomised Study Comparing First-line Dual Vs Single-Stentretriever Technique: TWIN2WIN

**DOI:** 10.1101/2024.07.11.24310312

**Authors:** Alejandro Tomasello, Manuel Moreu, Mikel Terceño, Lavinia Dinia, Maria Rosario Barrena Caballo, Manuel Requena, Magda Jablonska, Judith Cendrero, Alan Flores, Santy Ortega, Francesco Diana, David Henandez, Marta de Dios, Marta Rubiera, Alvaro Garcia-Tornel, Federica Rizzo, Marta Olivé, Carlos Pérez-García, Carmen Trejo Gallego, Tomas Carmona, Marc Rodrigo-Gisbert, Carlos Molina, Marc Ribo

**Affiliations:** Interventional Neuroradiology, Vall d’Hebron University Hospital, Barcelona, Spain; Stroke Research Group, Vall d’Hebron Research Institute, Barcelona, Spain; Interventional Neuroradiology, Hospital Clínico San Carlos, Madrid, Spain; Stroke Unit, Department of Neurology, Hospital Universitari Dr. Josep Trueta, Girona, Spain; Radiology, Hospital de la Santa Creu i Sant Pau, Interventional Neuroradiology Section, Barcelona, Spain; Interventional Neuroradiology Unit. Hospital Universitario Miguel Servet. Grupo de Investigación en Neurociencias. Aragon Institute for Health Research (IIS Aragón), Zaragoza, Spain; Stroke Unit. Neurology Department. Hospital Vall d’Hebron. Barcelona. Spain; Stroke Unit. Vall d’Hebron institut de Recerca. Barcelona. Spain; Neurology department. Hospital Joan XXIII. Tarragona. Spain; Department of Neurology, Neurosurgery and Radiology. University of Iowa. Iowa City. US; Dipartimento di Scienze della Vita, della Salute e delle Professioni Sanitarie, Università degli Studi, Rome, Italy; Neurosurgery Department, Hospital San Pablo, Coquimbo, Chile

## Abstract

**Importance:** Double stentretriever (double-SR) is used as a rescue technique when recanalization is not achieved in stroke patients undergoing thrombectomy. Double-SR, if applied as first-line technique could increase first-pass recanalization rates, known to be associated with better outcomes.

**Objective:** To assess the safety and efficacy of first-line double-SR in stroke patients undergoing thrombectomy.

**Design:** Randomized, controlled, blinded adjudicated primary outcome study between 2022 and 2023.

**Setting:** Multicenter (5 sites), national (Spain).

**Participants:** Patients with ischemic stroke due to large vessel occlusion within 24 hours after onset, undergoing thrombectomy.

**Interventions:** Upon confirmation of large vessel occlusion on initial angiogram, patients were randomly allocated to receive a first-line strategy: single-SR Vs double-SR technique. Investigators could use their technique of choice if further passes were needed.

**Main Outcomes and Measures:** The primary objective was to evaluate the efficacy of double-SR defined as first-pass complete recanalization (eTICI 2c-3) compared to single-SR. First/pass and final successful recanalization (eTICI2b50-3) were centrally assessed by a blinded investigator. The safety outcome was the occurrence of a symptomatic intracerebral hemorrhage (sICH). The data safety monitoring board stopped the recruitment after a pre-planned interim analysis because a predefined efficacy boundary was reached.

**Results:** From April 2022 to October 2023, 108 patients were included, 50 patients (46%) in the single-SR group and 58 (54%) in the double-SR group. FPR was achieved in 12/50 patients (24%) allocated to single-SR and 27/58 (46%) allocated to double-SR (aOR 2.72; 95% CI, 1.19-6.46). Substantial reperfusion within 3 attempts was obtained in 42 patients (84%) allocated to single-SR and in 52 (89%) allocated to double-SR (aOR 1.74; 95% CI, 0.55 - 5.76). The mean number of passes was 2±1.3 with single-SR and 1.7±1 with double-SR (mean difference, −0.37; 95% CI, −0.79 - 0.06). A sICH occurred in 3 patients (6%) allocated to single-SR and in 6 (10%) allocated to double-SR (aOR 1.66; 95% CI, 0.40-8.35).

**Conclusions and Relevance:** In stroke patients undergoing thrombectomy, first-line double-SR is safe and superior to single-SR in achieving first pass but not final recanalization. Implications on clinical outcomes should be studied in specifically designed trials.

**Trial Identification:** NCT05632458

**Key points:** *Question:* What is the safety and efficacy of the double stentretriever (SR) technique as a first-line treatment in acute ischemic stroke patients undergoing endovascular treatment?

*Findings:* In this multicenter randomized, blinded primary endpoint adjudicated clinical trial that included 108 acute stroke patients, the rate of first-pass recanalization (TICI2c-3) was superior with double-SR as compared to single-SR technique.

*Meaning:* In acute stroke patients with a large vessel occlusion, the first-line use of the double SR technique increases the chances of first-pass recanalization, which has been associated with improved clinical outcomes.

## INTRODUCTION

Reperfusion treatments have substantially and progressively improved acute stroke care over the last two decades by increasing recanalization outcomes. Tissue plasminogen activator has been the only approved specific treatment for acute ischemic stroke from 1995^1^ until 2015^2^ when most clinical guidelines also recommended mechanical thrombectomy as a first-line treatment in ischemic strokes due to a large vessel occlusion (LVO). In recent years, the number of patients benefiting from endovascular treatment (EVT) has rapidly grown worldwide and its indications progressively expanded^3,4,5^.

ALthough EVT has shown to consistently achieve substantial recanalization in 85-90% of the cases, more than 50% of treated patients will develop a moderate to severe disability, indicating substantial room for improvement^6^. Studies have shown that achieving complete recanalization (modified Thrombolysis in Cerebral Infarction (eTICI) 2c–3) at the first attempt (first pass recanalization (FPR)) is a strong predictor of functional independence (modified Rankin Scale score at 90 days 0–2)^7^ with each additional attempt progressively decreasing the probability of favorable outcome^8^. Furthermore, achieving a sudden recanalization—that is, complete clot retrieval with no clot fragmentation in a single pass, has also been associated with improved functional outcomes^9^.

Unfortunately, currently available thrombectomy devices to date can only achieve FPR rates ranging from 30 to 60%^10,11,12^. The simultaneous double stent-retriever (double-SR) technique was first proposed as a rescue therapy after multiple failed attempts to retrieve a refractory thrombus with a single-SR^13,14^. Recently, to maximize the rates of FPR, the double-SR technique has been proposed as the first-line treatment, and initial experiences have shown promising safety and efficacy results^15^. In this randomized clinical trial, we aim to investigate the safety and efficacy of the first-line double-SR technique as compared with the traditional single-SR approach.

## METHODS

### Study design and participants

TWIN2WIN is a randomized, multicenter, blinded adjudicated outcome, Phase II Clinical Study, designed to assess if the first-line use of the double-SR technique in stroke patients with confirmed LVO undergoing EVT is safe and effective as compared to the single-SR approach. This study followed the Consolidated Standards of Reporting Trials (CONSORT) reporting guideline. The trial was performed at 5 comprehensive stroke centers in Spain. The study was approved by the ethics committee at each site. Signed informed consent was obtained from the patients or their legally authorized representative. The study protocol is available in Appendix 1 (Supplemental material). The trial was designed and conducted by independent academic investigators, without the collaboration of a study sponsor. The primary outcome and other neuroimaging-related assessments were performed by two blinded independent expert readers after all imaging was uploaded to a central core laboratory. All clinical investigational activities were in accordance with the Clinical Investigation Protocol and Good Clinical Practices. An independent data safety monitoring board (DSMB) provided additional oversight of the study conduct.

Eligible patients were men and women with a disabling ischemic stroke at the time of randomization (baseline National Institutes of Health Stroke Scale [NIHSS] ≥6; range 0–42, with higher scores indicating greater stroke severity)^16^; functionally independent before the stroke, defined as a modified Rankin Scale (mRS) score between 0 and 2, with scores ranging from 0 (no symptoms) to 6 (death)^17^; in which EVT could be initiated within 24 hours from symptom onset or last time seen well.

Patients with a single large vessel occlusion at the level of the terminal internal carotid artery (ICA), the M1 segment of the middle cerebral artery (M1-MCA) or the top of the basilar artery on initial digital substraction angiogram of the EVT, were evaluated as candidates for the TWIN2WIN study. No screening log was kept. A full list of eligibility criteria is available in Appendix 2 (Supplemental material).

### Enrollment, randomization and masking

On confirmation of eligibility, patients were randomly allocated 1:1 using the studyrandomizer web-based blocked dynamic stratified randomization tool to be treated in the first thrombectomy attempt with the double-SR or the single-SR technique (Appendix 3, Supplemental). A fixed, nonadaptive, randomization scheme was used. Stratification was completed by center and occlusion site on baseline angiogram: terminal ICA, M1-MCA, or basilar artery occlusion. Admission neuroimaging, angiographic runs, and follow-up CT scans were recorded and anonymized for subsequent blinded outcome assessment in a central imaging core lab by experienced neurointerventionalists blinded to the allocated study arm. The quality of reperfusion was assessed with the Expanded Treatment in Cerebral Infarction [eTICI] scale^6^ (range of 0 to 3, with higher grades indicating increased reperfusion).

### Procedures

Upon arrival at the hospital patients were treated according to local institutional protocols, and national and international guidelines including intravenous thrombolysis when indicated^18^. The arterial puncture was performed within 90 minutes of the mandatory screening CT or magnetic resonance imaging. After qualifying baseline angiogram, all patients intended to receive EVT were immediately included and randomized. Decisions regarding the type of anesthesia and use of balloon-guided catheters were at the discretion of the operators, the concomitant use of a distal access catheter (DAC) was suggested.

Per protocol, the first pass technique was determined by the allocated study arm. In the single-SR approach, the combination therapy with a distal access catheter (DAC) was typically used: a 0.021’’ microcatheter was advanced through the occlusion to a distal branch, and the SR was deployed. The thrombectomy pass was then performed according to the neurointerventionalist choice (complete Vs partial SR retrieval into the DAC^19^). In the double-SR technique, a second microcatheter was placed distal to the occlusion before SR deployment, preferably in a different branch (Y configuration); that is, the A1-ACA and M1-MCA for terminal ICA occlusions, 2 different M2-MCA branches for M1-MCA occlusions and each P1-PCA segment for terminal basilar artery occlusions. Whenever 2 different distal branches could not be easily catheterized, the interventionalist could place both microcatheters in parallel in the same branch distal to the occlusion. After placing both microcatheters in the target vessels, the microwires were exchanged for the SRs (any combination of commercially available SRs was allowed), which were then unsheathed across the clot by withdrawing the microcatheters. Typically, the DAC was then navigated to the proximal interface of the thrombus, the microcatheters were removed from the stent retriever pusher wires to maximize the aspiration efficacy in the DAC^20^. Finally, the SRs were partially (approximately one-third) and simultaneously retrieved into the DAC under continuous aspiration before the whole system was pulled into the guiding catheter. (Appendix 6 Supplemental material)

If, after the first pass, thrombectomy with the allocated technique failed to restore near complete cerebral blood flow (eTICI grade 2c to 3 [grade 2c indicates reperfusion of ≥90% and grade 3 100% of the initially affected territory], the patient was assigned as technique failure for the primary endpoint. In these cases, the treating physician could decide what treatment therapy (if any) was deemed appropriate for the patient. If additional thrombectomy passes were performed after the first pass, the total number of passes, the procedural duration, and the final eTICI score were recorded.

Sites were expected to adhere to national guidelines for stroke units, stroke rehabilitation, and stroke prevention care^21^. All patients had standard assessments of demographic characteristics, past medical history, laboratory values, and stroke severity (NIHSS score). Admission imaging consisted at least of a non-contrast CT and CT-angiography CT-perfusion was performed when considered necessary by treating physicians. Follow-up non-contrast CT was performed at 24h.

Following the index procedure, each patient was subjected to a follow-up assessment at 24 hours (NIHSS), and 5 days or discharge (NIHSS + mRS). Clinical follow-up was also blindly obtained at 90 days from randomization, if possible in-person, to assess the degree of disability as determined by the mRS. Where in-person follow-up was not possible, video conferencing or telephone follow-up was obtained^22^. (eAppendix 4 in Supplemental)

### Efficacy Outcomes

The objective of the study was to evaluate the efficacy of the first-line double-SR technique compared to the single-SR approach. The primary efficacy endpoint was defined as the rate of first-pass angiographic complete recanalization (eTICI 2c-3). Secondary efficacy endpoints included: 1) successful reperfusion, defined as the achievement of a modified Cerebral Infarct Thrombolysis eTICI2b50 or greater in the target vessel after 3 passes, 2) workflow times, including time from onset to reperfusion (if achieved) and time from groin puncture to reperfusion (if achieved), 3) number of retrieval or aspiration attempts performed during the procedure 4) neurological status at 24 hours determined by the NIHSS score, 4) mRS score at discharge or 5 days (early assessment) and at 90 days (late assessment), dichotomized into mRs score of 0-1 and 2-6.

### Safety Outcomes

The primary safety endpoint was the rate of symptomatic intracerebral hemorrhage (sICH) within 24 hours (range of 16 to 36 hours) postprocedure according to the Safe Implementation of Thrombolysis in Stroke-Monitoring Study (SITS-MOST) criteria^23^ (local or remote parenchymal hemorrhage type 2 on the posttreatment imaging combined with a neurologic deterioration of 4 points or more compared with baseline or the lowest posttreatment NIHSS value or death). Secondary safety endpoints included: 1) any intracerebral hemorrhage at 24 hours, 2) neurological deterioration defined as ≥4 points on the NIHSS at 24 hours assessed by an investigator not involved in the thrombectomy procedure, 3) embolization in a previously uninvolved territory in the final cerebral angiogram, 4) procedure-related mortality rate at 5 days (+/- 12h) or at discharge, 5) procedural complications (arterial perforation, arterial dissection).

### Statistical analyses, sample size calculation, and early termination

The reported first-pass reperfusion for proximal vessel occlusions among stroke patients undergoing EVT with single-SR ranges from 33-50%. We estimate that double-SR could increase the first-pass effect to 58%. With this assumption, to reach a statistical power of 75% to detect differences using a bilateral Chi-square test with a significance level of 5% and a 1/1 distribution per study arm, the required sample size is 212 patients (106 per study arm).

The primary analysis was performed based on the population as randomized. Patients who did not receive the allocated study treatment were included in the primary analysis within the respective treatment group they had been randomized. A designated Data Safety Monitoring Board (DSMB) planned to review the primary safety endpoint after every 25 new patients enrolled in the study and determine study continuation according to the occurrence of adverse events in each arm of the study. A single interim efficacy analysis was planned to be performed after the inclusion of 106 patients (50% of the total sample size) and recruitment stopped if an overwhelming efficacy in first-pass recanalization is demonstrated between the two groups according to pre-specified stopping rules based on the Pocock boundary method^24^. (Appendix 5, supplemental). The analysis of the primary and secondary endpoint analyses was based on the observed data. There were no missing values in the primary outcome or main secondary outcomes. In one patient the 90-day mRS could not be assessed and the last observation (mRS at discharge) was carried forward to impute the missing value.

The primary and secondary outcomes that assessed binary dependent variables were estimated based on a logistic regression model adjusted by initial occlusion location (TICA, MCA-M1, and MCA-M2) and described as odds ratio and 95% confidence intervals. Between-group differences in continuous outcomes were assessed based on a t-test, expressed as the mean difference and 95% CI. A preplanned interim analysis based on Pocock stopping rules assessed the primary outcome in 108 patients, considering evidence of an overwhelming efficacy a p-value of 0.025 or less in the between-group difference in the rate of FPR. All secondary outcomes are reported with effect measures and 95% CI, without adjustment for multiple comparisons. All statistical analyses were performed using the R 4.3.3 version (R project for statistical computing).

## RESULTS

Between April 2022 and November 2023, a total of 108 patients were randomized across 5 sites. Of these, 55 (50.9%) were female, and the median [IQR] age was 77 [69-82.5] years. A total of 50 patients (46.3%) were randomized to the single-SR group and 58 (53.7%) to the double-SR group. The study-allocated procedure was performed on the 107 patients (99.1%, as treated population), as one patient allocated to the double-SR technique was treated with single-SR. Baseline demographic, clinical, and imaging characteristics were comparable between the 2 trial groups (Table 1). The median [IQR] baseline NIHSS score was 18 [14-21] and the median [IQR] ASPECTS was 9 [7-10]. A total of 38 patients (35.2%) received intravenous thrombolysis. Occlusion locations were TICA in 37 patients (34.3%), M1-MCA in 63 patients (58.3%), and basilar in 8 patients (7.4%). The median time from stroke onset to arterial puncture was 387.6 [330.9 - 444.1] minutes, and the median procedural time was 44.9 [39.6 - 50.2] minutes.

**Table 1.**
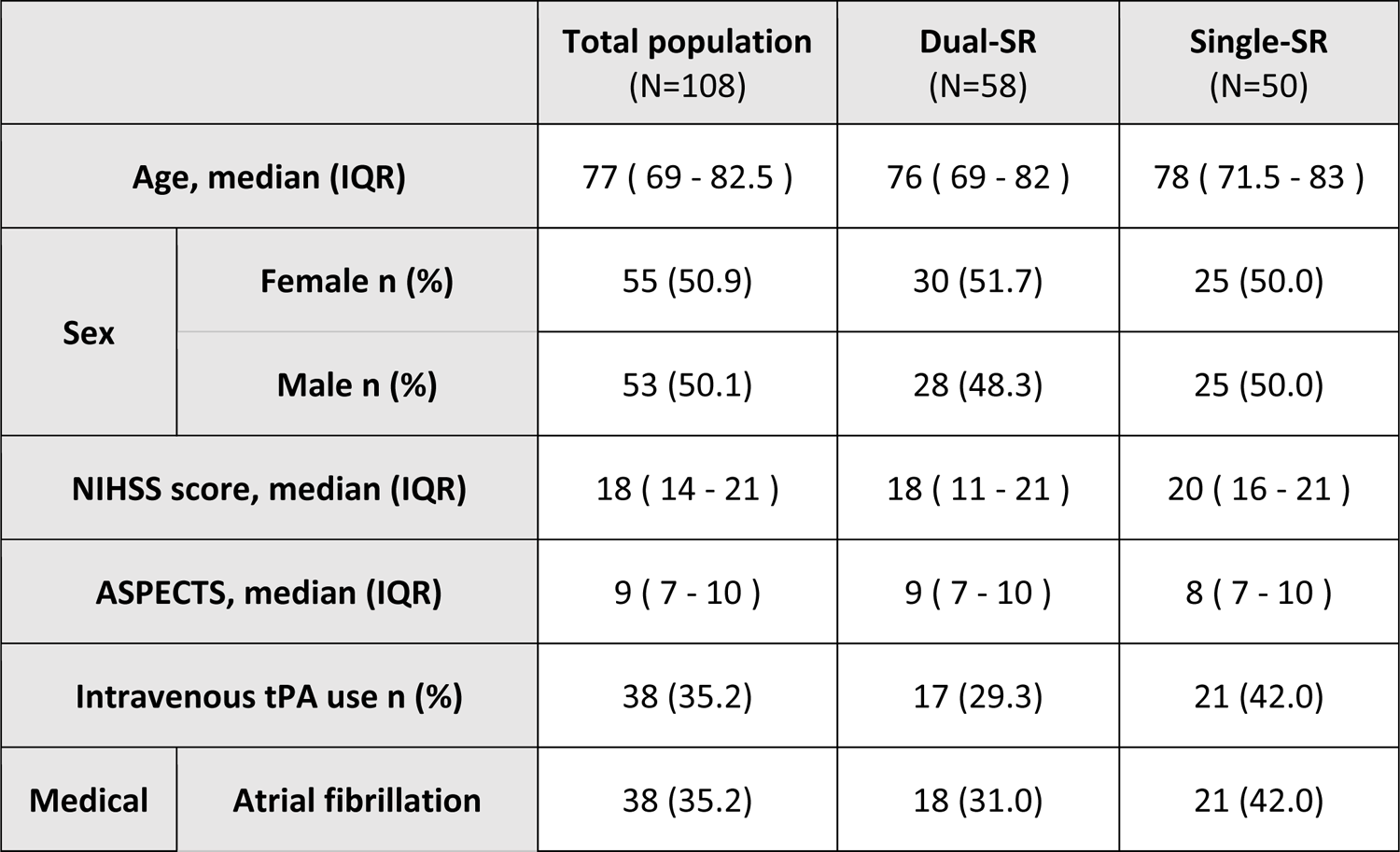
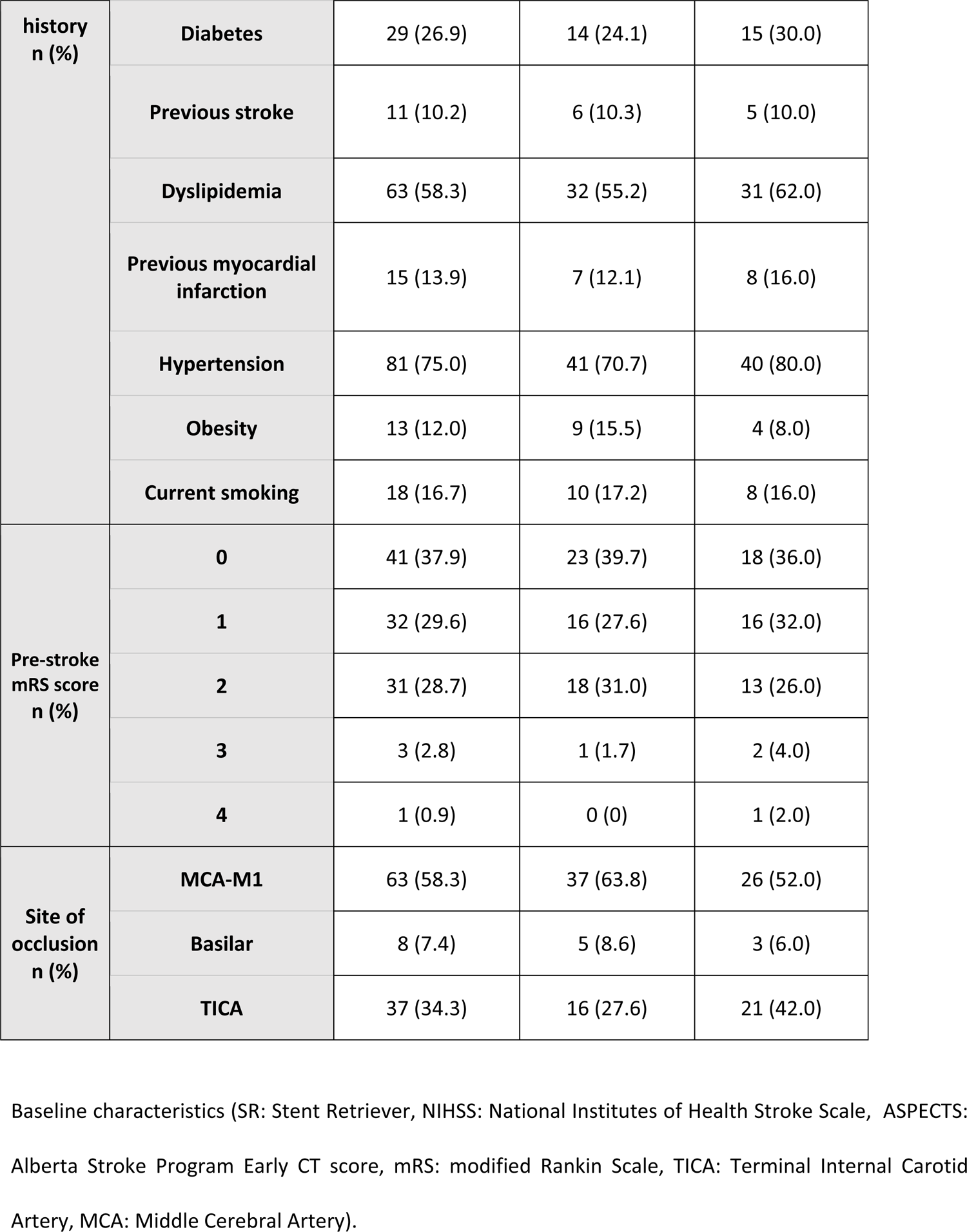
Baseline characteristics.

|  |  | **Total population**<br>**(N=108)** | **Dual-SR**<br>**(N=58)** | **Single-SR**<br>**(N=50)** |
| --- | --- | --- | --- | --- |
| **Age, median (IQR)** |  | 77 ( 69 - 82.5 ) | 76 ( 69 - 82 ) | 78 ( 71.5 - 83 ) |
| **Sex** | **Female n (%)** | 55 (50.9) | 30 (51.7) | 25 (50.0) |
|  | **Male n (%)** | 53 (50.1) | 28 (48.3) | 25 (50.0) |
| **NIHSS score, median (IQR)** |  | 18 ( 14 - 21 ) | 18 ( 11 - 21 ) | 20 ( 16 - 21 ) |
| **ASPECTS, median (IQR)** |  | 9 ( 7 - 10 ) | 9 ( 7 - 10 ) | 8 ( 7 - 10 ) |
| **Intravenous tPA use n (%)** |  | 38 (35.2) | 17 (29.3) | 21 (42.0) |
| **Medical** | **Atrial fibrillation** | 38 (35.2) | 18 (31.0) | 21 (42.0) |
| **history**<br>**n (%)** | **Diabetes** | 29 (26.9) | 14 (24.1) | 15 (30.0) |
|  | **Previous stroke** | 11 (10.2) | 6 (10.3) | 5 (10.0) |
|  | **Dyslipidemia** | 63 (58.3) | 32 (55.2) | 31 (62.0) |
|  | **Previous myocardial infarction** | 15 (13.9) | 7 (12.1) | 8 (16.0) |
|  | **Hypertension** | 81 (75.0) | 41 (70.7) | 40 (80.0) |
|  | **Obesity** | 13 (12.0) | 9 (15.5) | 4 (8.0) |
|  | **Current smoking** | 18 (16.7) | 10 (17.2) | 8 (16.0) |
| **Pre-stroke**<br>**mRS score**<br>**n (%)** | **0** | 41 (37.9) | 23 (39.7) | 18 (36.0) |
|  | **1** | 32 (29.6) | 16 (27.6) | 16 (32.0) |
|  | **2** | 31 (28.7) | 18 (31.0) | 13 (26.0) |
|  | **3** | 3 (2.8) | 1 (1.7) | 2 (4.0) |
|  | **4** | 1 (0.9) | 0 (0) | 1 (2.0) |
| **Site of**<br>**occlusion**<br>**n (%)** | **MCA-M1** | 63 (58.3) | 37 (63.8) | 26 (52.0) |
|  | **Basilar** | 8 (7.4) | 5 (8.6) | 3 (6.0) |
|  | **TICA** | 37 (34.3) | 16 (27.6) | 21 (42.0) |

Baseline characteristics (SR: Stent Retriever, NIHSS: National Institutes of Health Stroke Scale, ASPECTS: Alberta Stroke Program Early CT score, mRS: modified Rankin Scale, TICA: Terminal Internal Carotid Artery, MCA: Middle Cerebral Artery).

The DSMB did not find safety concerns in the interim safety analysis (Appendix 5, Supplemental). On the preplanned efficacy analysis, after 108 patients were enrolled, the DSMB recommended stopping recruitment because the predefined efficacy boundary was reached (p = 0.019).

### Efficacy Outcomes

The primary efficacy outcome, the rate of FPR among included patients in the population as-randomized, was 12/50 patients (24%) in the single-SR group and 27/58 patients (47%) in the double SR group (aOR 2.72; 95% CI, 1.19 to 6.46, p=0.019). Successful reperfusion within 3 thrombectomy attempts was achieved in 42/50 patients (84%) in the single-SR group and 52/58 patients (89%) in the double-SR group (aOR 1.74, 95% CI 0.55 to 5.75). At the end of the procedure, successful reperfusion was achieved in 48/50 patients (96%) in the single-SR group and 43/58 patients (93%) in the double-SR group (aOR 0.55, 95% CI 0.07 to 3.01)

Time from stroke onset to groin puncture was 274 minutes (IQR 185-520) in the single-SR group and 292 minutes (IQR 207-275) in the double-SR group (mean difference: −1 minute, 95% CI-114 to 113). Time from groin puncture to reperfusion was 36 minutes (IQR 27-57) in the single-SR group and 39 minutes (IQR 27-63) in the double-SR group (mean difference: 6 minutes 95% CI-4 to 16).. A mRS score of 0-1 at 90 days was achieved in 7/50 patients (14%) in the single-SR group and 16/58 patients (28%) in the double-SR group (aOR 2.31, 95% CI 0.88 to 6.69). All secondary efficacy outcomes are presented in Table 2

**Table 2:** Efficacy and safety outcomes (SR: Stent Retriever, eTICI: expanded Thrombolysis in Cerebral Ischemia, NIHSS: National Institutes of Health Stroke Scale, mRS: modified Rankin Scale, sICH: symptomatic Intracerebral Hemorrhage).

|  | **Single-SR**<br>**(N=50)** | **Dual-SR**<br>**(N=58)** | **Treatment effect**<br>**[IC - 95%]** | **Effect measure** |
| --- | --- | --- | --- | --- |
| **Primary efficacy outcome**<br>First-pass complete<br>recanalization (eTICI 2C-3) | 12 (24%) | 27 (47%) | 2.72 (1.19 to 6.46) | Odds ratio |
| **Secondary efficacy outcome**<br>Successful reperfusion<br>within 3 passes (eTICI 2B-3) | 42 (84%) | 52 (89%) | 1.74 (0.55 to 5.75) | Odds ratio |
| Time from groin puncture to<br>reperfusion<br>Median (IQR) | 36 (27-57) | 39 (27-63) | 6 (-4 to 16) | Mean difference |
| Number of passes<br>Median (IQR) | 3 (2-3) | 2 (2-3) | -0.37 (-0.79 to 0.06) | Mean difference |
| NIHSS at 24 hours<br>Median(IQR) | 13 (4-20) | 10 (3-20) | -1.1 (-4.9 to 2.6) | Mean difference |
| mRs 0-1 at 5 days<br>or discharge | 9 (18%) | 12 (20%) | 1.18 (0.45 to 3.20) | Odds ratio |
| mRs 0-1 at 90 days | 7 (14%) | 16 (28%) | 2.31 (0.88 to 6.69) | Odds ratio |
| **Primary safety outcome**<br>Symptomatic intracerebral<br>haemorrhage (SICHs) | 3 (6%) | 6 (10%) | 1.66 (0.40 to 8.35) | Odds ratio |
| **Secondary safety outcome**<br>Intracerebral haemorrhage | 10 (20%) | 18 (31%) | 1.78 (0.73 to 4.51) | Odds ratio |
| Neurological deterioration<br>(≥4 points on NIHSS) at 24h | 6 (12%) | 9 (16%) | 1.15 (0.37 to 3.76) | Odds ratio |
| Embolization in a previously<br>uninvolved territory | 1 (2%) | 1 (2%) | 0.96 (0.03 to 25.38) | Odds ratio |
| Procedure-related mortality at<br>5 days or discharge | 1 (2%) | 2 (3%) | 1.78 (0.16 to 39.73) | Odds ratio |
| Procedure complications | 3 (6%) | 5 (9%) | 1.38 (0.32 to 7.15) | Odds ratio |

### Safety Outcomes

The primary safety outcome, the occurrence of sICH, was reported in 3 patients (6%) in the single-SR group and in 6 patients (10%) in the double-SR group (aOR 1.66, 95% CI 0.40 to 8.35). Any intracranial hemorrhage after the procedure was diagnosed in 10 patients (20%) in the single-SR group and 18 patients (31%) in the double-SR group (aOR 1.78, 95% CI 0.73 to 4.51). The number of patients who experienced complications of the procedure was 3 (6%) in the single-SR group and 5 (9%) in the double-SR group (aOR 1.38, 95% CI 0.32 to 7.15). All secondary safety outcomes are presented in Table 2. A descriptive analysis of all adverse events is presented (Table 3)

**Table 3:** adverse events and serious adverse events (SR: Stent Retriever)

|  | **Single-SR**<br>**(N=50)** | **Dual-SR**<br>**(N=58)** |
| --- | --- | --- |
| **Adverse Events** |  |  |
| Intracranial hemorrhage | 10 (20%) | 18 (31%) |
| Thrombosis | 0 (0%) | 1 (2%) |
| Infection | 1 (2%) | 1 (2%) |
| Others | 1 (2%) | 3 (5%) |
| **Serious Adverse Events** |  |  |
| Stroke progression | 1 (2%) | 7 (12%) |
| Intracranial hemorrhage | 3 (6%) | 6 (10%) |
| Malignant stroke | 3 (6%) | 3 (5%) |
| Pulmonary embolism | 1 (2%) | 0 (0%) |
| Infection | 1 (2%) | 0 (0%) |

## DISCUSSION

This randomized clinical study demonstrates that the first-line double-SR technique is more effective than single-SR in achieving FPR when treating proximal large vessel occlusions in acute stroke patients undergoing EVT. Additionally, the study did not show evident safety concerns related to the double-SR technique, however, the study was not powered to confirm this extent. Since a lower number of thrombectomy attempts^8^ and particularly FPR^7^, have been repeatedly shown to be associated with improved clinical outcomes, the results of this study open the door to consider the double-SR, at least in selected cases, as the standard first-line approach instead of a rescue technique to be applied only when multiples attempts had failed to induce recanalization. Further studies confirming the clinical benefit, safety, and potential optimal occlusion locations such as vessel bifurcation are however warranted before formal recommendations can be made.

To date, multiple case series have reported the use of the double-SR technique as a valuable bail-out approach when conventional techniques had failed^13,14^. A recent in-vitro study demonstrated the benefits of the double-SR technique in occlusions occurring at the level of vessel bifurcations, where saddle clots are switched from one distal branch to the other with each additional single-SR pass^14^. Placing a SR in each of the distal segments avoids rolling of the clot into the spare distal vessel branch during retrieval increasing the chances of success^25^. A different study also showed how the clot-to-vessel contact area is dramatically reduced when 2 SRs are deployed sandwiching the clot as compared to a single SR^26^. Limiting the clot-vessel wall contact area reduces the friction experienced by the clot during retrieval and therefore the chances of clot roll-out and loss during the maneuver. To ensure that both stents are separately and independently deployed it is essential to cross the clot with two microcatheters before simultaneous deployment. Otherwise, in a sequential delivery, after deploying the first SR, the second microcatheter will most probably advance coaxially inside the first SR reducing the chances of “sandwiching” the clot between both SRs. Moreover, the presence of 2 separated SR ensures an increased radial force of the devices against the clot through all the thrombectomy passes despite the progressive increase in the vessel diameter when approaching the guiding catheter in the ICA. Once the SRs deployed, the DAC was suggested to be advanced until approximately one-third of the SRS are inside the DAC, acting like a zipper that pinches the clot before retrieval (figure Appendix 6, supplemental material). The most effective double-SR technique however still needs to be determined, but will probably vary according to biomechanical clot characteristics which at present cannot be accurately determined before retrieval.

The study showed the superiority of first-line double-SR in achieving complete recanalization at the first pass but not after 3 passes or at the end of the procedure, where both arms showed comparable results. However, in order to achieve the same degree of final recanalization, patients in the single-SR arm needed to undergo more passes which is a factor known to negatively impact clinical outcome^8^. Moreover, the study protocol allowed technique crossover after the first pass, and several patients initially allocated to the single-SR arm were treated with double-SR as a rescue technique (3/50, 6%), which may have contributed to homogenize the rates of final recanalization.

The TWIN2WIN study raises some concerns about the safety of the double-SR technique, as the rate of sICH was numerically higher in the double-SR arm. Despite that, the mortality rates were similar in both groups. A previous animal study showed a moderate increase in endothelial damage induced by double-SR as compared to single-SR technique^27^. In the present study, the incidence of adverse events in the double-SR arm was not statistically higher than in the single-SR arm and comparable to the rates reported in previous EVT studies^2^. However, the trial was not powered to detect potential differences in safety outcomes and future trials will need to specifically address the safety profile of the technique.

The present pilot study was not powered to determine the impact of the double-SR technique on the clinical outcome, and first-pass recanalization was used as a surrogate outcome measure. Future studies should investigate if the preliminary findings of numerically higher rates of excellent outcomes (mRS 0-1) observed in the double-SR arm can be replicated and robustly confirmed.

A frequently alleged reason to not adopt the double-SR from the first thrombectomy pass is the impact on procedural costs. Systematically adopting the double-SR as the frontline approach may increase the procedural complexity and cost of treatment compared with other commonly used techniques. However, under specific anatomical conditions, it can decrease the number of device passes or rescue strategies required to achieve revascularization, improving clinical outcomes and reducing the economic burden of post-stroke care. If the clinical benefits of the double-SR technique are confirmed, specific cost-effective studies should address this issue.

Our study has some limitations that are mainly related to budgetary restrictions. TWIN2WIN was an investigator-initiated study completed without any specific industry or public funding. The clinical data, which only included information usually prospectively collected in the institutional stroke registries, were not monitored by an independent clinical research organization. However, the primary endpoint was blindly adjudicated by an independent central core laboratory. The rate of first-pass recanalization observed in the single-SR arm might seem low compared to previously reported series^11^. The fact that in TWIN2WIN the initial occlusion was often located at the level of a bifurcation (terminal ICA, distal MCA, or top of basilar artery) has probably contributed to these lower than initially expected FPR rates. In any case, patients were randomly allocated and received the treatment after confirmation of the occlusion site on the initial angiogram. The improved efficacy of the double-SR technique may vary among different occlusion locations, particularly in vessel bifurcation occlusions when clots are switched from one branch to the other but not retrieved with each additional single-SR pass^14^.

## CONCLUSIONS

In stroke patients undergoing EVT, first-line double-SR is superior to single-SR in achieving first pass but not final recanalization. Implications on safety and clinical outcomes should be further studied in specifically designed trials.

**Trial Identification**: NCT05632458

## Disclosures

Marc Ribo is the co-founder of Anaconda Biomed and Norahealth and has a consulting agreement with Medtronic, Johnson&Johnson, Vesalio, Stryker, Philips, Apta Targets.

**Figure 1.**
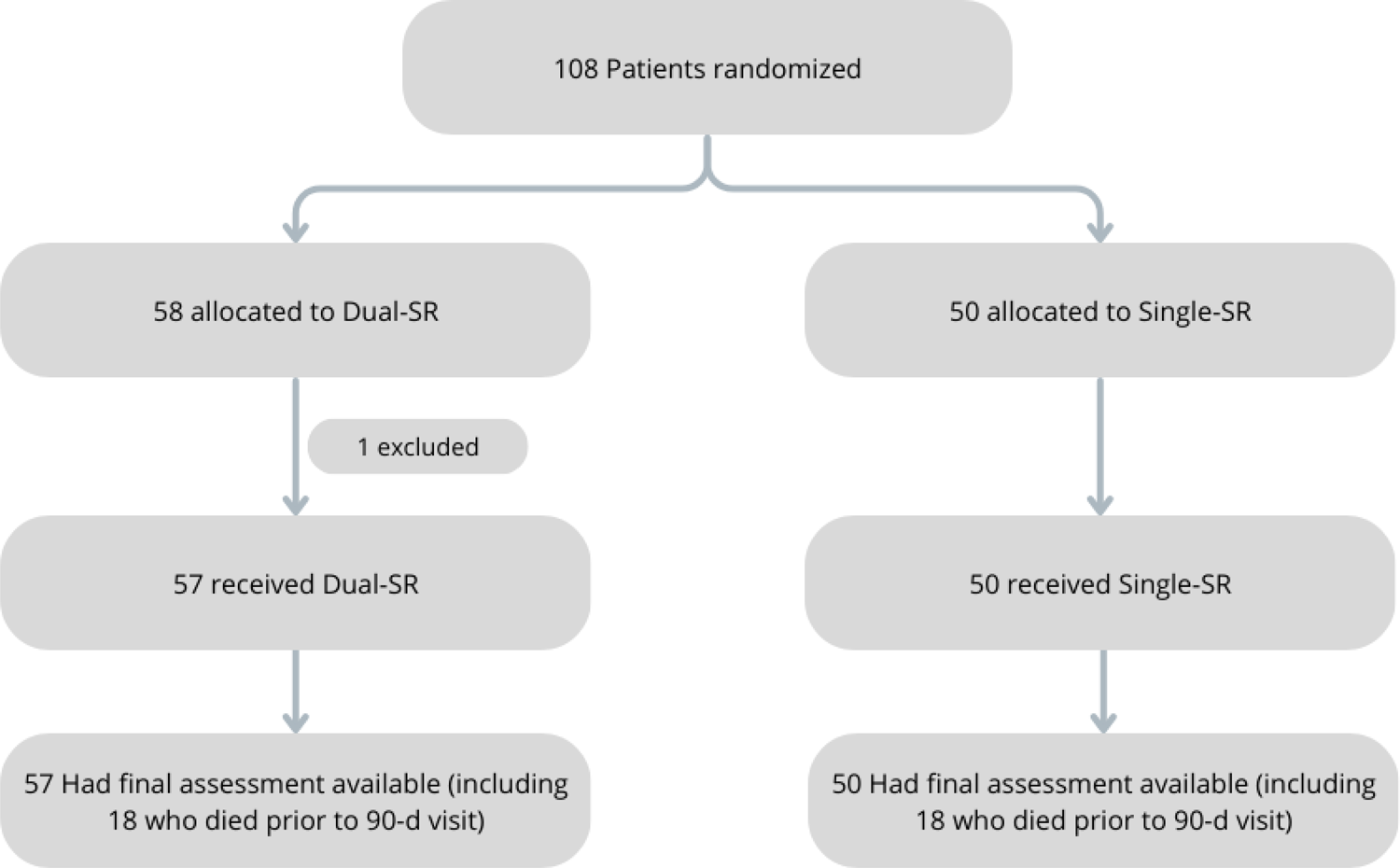
Study flowchart

**Figure 2.**
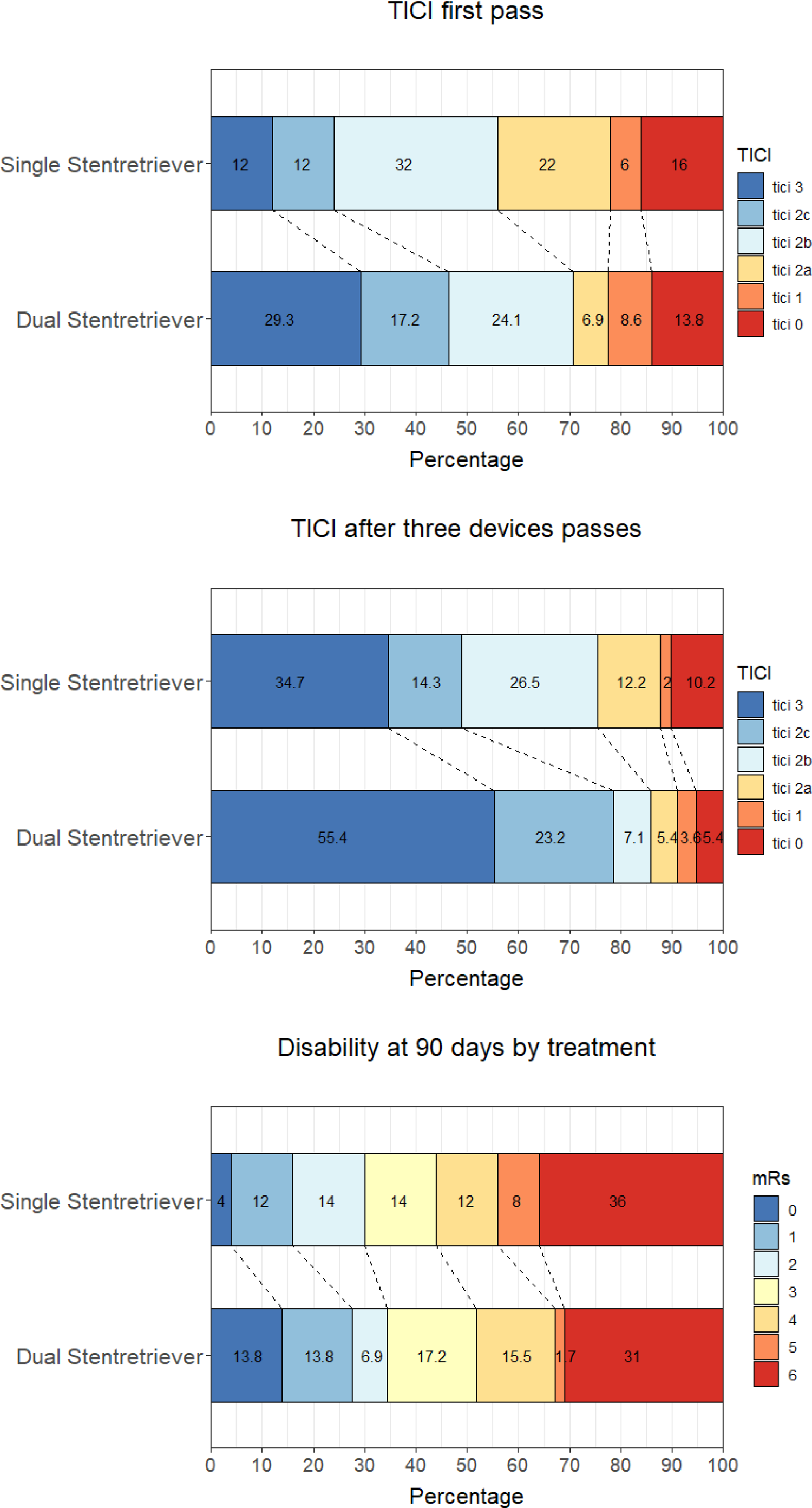
Primary and Secondary efficacy outcomes (TICI: expanded Thrombolysis in Cerebral Ischemia, mRS: modified Rankin Scale)

## Data Availability

Relevant data will be shared upon reasonable request from researchers with trackable record in the field.

